# Examining the governance arrangements for healthcare worker COVID-19 protection in Kenya: A Scoping Review

**DOI:** 10.1101/2023.02.22.23286303

**Authors:** Jacob Kazungu, Nancy N Kagwanja, Huihui Wang, Jane Chuma, Kenneth Munge

## Abstract

Healthcare workers (HCWs) face a high risk of infection during pandemics or public health emergencies as demonstrated in the ongoing COVID-19 pandemic. Understanding how governments respond can inform public health control measures and support health system functioning. An economic impact analysis examining HCW COVID-19 infections in Kenya and three other countries estimated that the total economic costs related to HCW COVID-19 infections costs and deaths in Kenya were US$113.2 million (range US$35.8-US$246.1). We examined the governance arrangements for HCW protection during the COVID-19 pandemic in Kenya between March 2020 and March 2021.

Governance arrangements were examined following a scoping review of 44 policy and legislative documents and reports of HCW protection and 22 media articles. We adopted the transparency, accountability, participation, integrity and capacity (TAPIC) governance framework to analyse and summarize our findings into policy gaps and implementation challenges.

Policy design gaps included inadequate provisions for emerging threats, inconsistencies with the devolved context and inadequate structures to monitor, inform and respond to HCW COVID-19 infections. Implementation challenges were attributed to inadequate quantity and quality of PPE, difficulty in accessing medical care for HCWs, delays in HCW remuneration, insufficient infection prevention and control measures, the top-down application of plans, difficulties in working in a decentralized context, and pre-existing public finance management (PFM) bottlenecks.

Implementation of HCW protection during the COVID-19 pandemic and beyond could leverage the revamping of current legislation on labour relations to reflect devolved governance and develop a broader and long-term approach to occupational health and safety implementation that considers all HCWs. Improvements in PFM arrangements coupled with increased investment in the health sector and attention to efficient use of resources will also impact positively on HCW protection.

## INTRODUCTION

Healthcare workers (HCWs) are a key component of preparedness and response to public health emergencies including pandemics. The COVID-19 pandemic has highlighted that HCWs are, among other things, at high risk of exposure to infection, and other impacts of pandemics, due to their role in front-line service delivery. For instance, existing evidence indicated that HCWs were over 11 times more likely to test positive for COVID-19 compared to the general population (1). In low- and middle-income countries (LMICs) such as Kenya, pre-existing HCW challenges such as low staffing ratios (2), maldistribution, and general resource constraints can exacerbate these impacts (3).

As of November 30, 2021, a total of 7, 848 COVID-19 infections and 53 deaths among HCWs were reported in Kenya (4) (Figure 1). Every HCW death and infection due to COVID-19 is not only regrettable but also concerning given the economic impact related to treatment, transmission to others, absence from work by the affected HCWs, and compromised quality of care during the COVID-19 pandemic. An economic impact analysis examining HCW COVID-19 infections in Kenya and three other countries reported elsewhere estimated that the total economic costs related to HCW COVID-19 infections costs and deaths were US$113.2 million (range US$35.8-US$246.1) (5). The costs of infections of HCW, secondary infections to others and indirect costs due to maternal and child deaths were 4.6%, 13.2% and 82.2% respectively. The total cost was equivalent to 2% of total health expenditure.

**Figure 1:**
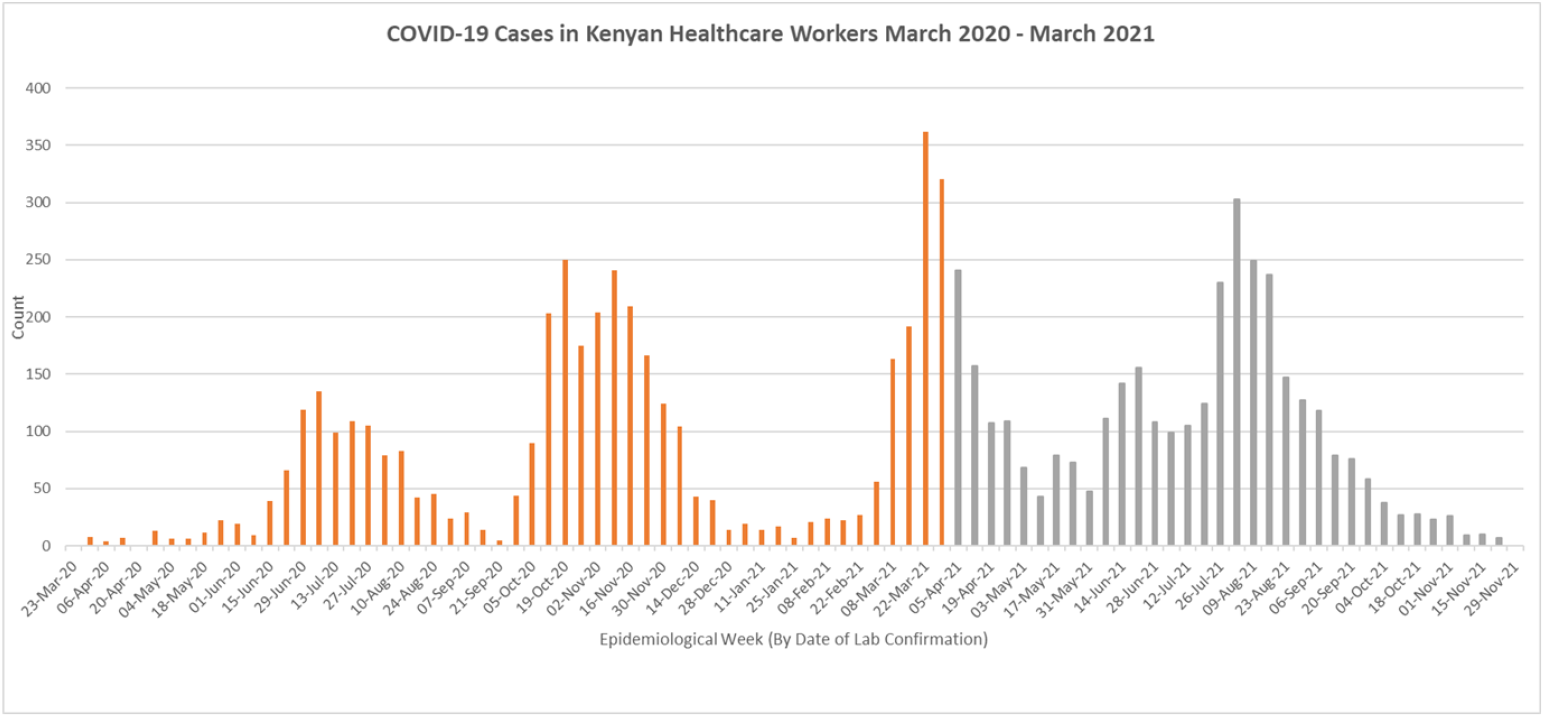
COVID-19 infection trends among HCWs as of November 30, 2021; Source: World Bank from MoH Kenya (2021), COVID-19 Outbreak in Kenya Daily Situational Report. *Orange bars indicate the study period*

This paper explores governance arrangements for HCW protection and implementation of COVID-19 HCW protections between March 2020 and March 2021. Specifically, we examined existing policy and legislative frameworkand implementation of HCW protection during the COVID-19 pandemic from a governance perspective. Brinkerhoff and Bossert (2008) describe governance as the rules determining the distribution and performance of roles and responsibilities among government, health service providers and beneficiaries (6). In this study, we draw on the transparency, accountability, participation, integrity and capacity (TAPIC) framework (Table 1) to examine how governance worked to achieve healthcare worker protections during the COVID-19 pandemic in Kenya (7). Specifically, we highlight the policy design gaps and implementation challenges identified across the TAPIC framework components.

**Table 1:** The components of the TAPIC framework (7).

| Component | Details |
| --- | --- |
| Transparency | Decisions, their grounds and their decision-makers are made clear. Access to public information for citizens is made clear. |
| Accountability | Process of holding individuals, agencies, and organisations (public, private, and civil society) responsible for executing their powers. |
| Participation | Engagement of affected parties/interests in decision-making |
| Integrity | Good public management such as meritocratic hiring, clear performance, standards, and clear organizational missions |
| Capacity | A mix of technical skills and networks, across governments and among experts and civil society. Also refers to the ability to formulate evidence-based and politically sensible recommendations for health |

Key terms used in this paper are summarised in Box 1:

### Box 1

Summary of key terms

- **Employer**: Any public body including national and county governments, and their agencies, or private organisation which has entered a contract of service to employ healthcare workers.
- **Healthcare worker (HCW)**: Everyone working in the healthcare sector and involved in the delivery of healthcare in any capacity, which includes but is not limited to doctors, nurses, laboratory staff, nutritionists, hospital cleaners, ambulance drivers, and administrative staff who support health service delivery.
- **Healthcare worker protection:** All the measures to promote the safety, welfare, and well-being of healthcare workers, and includes the ordinary basic or minimum wage or pay and emoluments payable, directly, or indirectly whether cash or in kind, by an employer to an employee arising out of the employment of that employee, medical benefits, provision of personal protective equipment (PPE) and a safe working environment. These protections are those that should be in place including during the COVID-19 pandemic and are drawn from Kenyan legislation and the guidelines on COVID-19 by WHO and ILO.

## METHODS

We conducted a scoping review of the governance arrangements and implementation of HCW protections for the COVID-19 response in Kenya. The documents related to proposed plans, reports and implementation of HCW protection were restricted to the period between March 2020 and March 2021 given the first officially reported case of COVID-19 in Kenya was recorded on March 13, 2020. However, we referred to documents published before March 2020 to examine existing governance arrangements for HCW protection before COVID-19.

### Scoping Review

To describe and analyse the Kenyan governance context for the protection of healthcare workers during the COVID-19 pandemic, we conducted a scoping review guided by the following questions:

1. What are the laws, policies and mechanisms that govern the protection of health workers in Kenya?
2. What are the roles and responsibilities of different levels of government and other institutions/stakeholders?
3. What are the decision-making, funding, oversight, monitoring and evaluation processes in place for HCW protection?
4. How and what provisions were made to enhance the protection of health workers during the COVID-19 pandemic?
5. To what extent did these provisions meet the needs of health workers during the COVID-19 pandemic, viewed through the lens of the TAPIC framework?

### Search strategy

We conducted purposive searches for articles and documents relevant to healthcare worker protection in two iterative phases (Table 2 summarises the search terms applied):

i. A search of the websites of various institutions to identify legislation, policy and guidelines on healthcare worker protection. These institutions included the Ministry of Health (MoH), Ministry of Labour and Social Protection (MoLSP), county government websites, World Health Organization (WHO), research institutions (e.g., Kenya Medical Research Institute (KEMRI)), civil society groups (e.g., the Kenya Legal and Ethical Issues Network (KELIN)), and HCW unions and associations. The same websites were searched for reports of incidents related to COVID-19 exposure among HCWs (for example infection rates, deaths, stigma) and practices of HCW protection during the COVID-19 pandemic.
ii. A search of media articles reporting on proposed and implemented HCW protection measures and on issues influencing the implementation of HCW protections during the COVID-19 pandemic period. This search was conducted on Google and the following search terms were applied.

**Table 2:** Search terms applied

| **Term**<br>**Outbreak**<br>Variants combined by OR | **Protection**<br>Variants combined by OR | **Health care worker**<br>Variants combined by OR |
| --- | --- | --- |
| COVID-19, coronavirus, SARS-CoV-2 pandemic, emergency, outbreak, epidemic | Protection, Intervention*, Infection, stress, burn-out, support, safety, mental health | Health provider, nurse* doctor*, clinician*, healthcare staff, healthcare provider* |

### Inclusion criteria

The following inclusion criteria were applied to the documents and articles.

1. Kenyan legislation and policy documents related to healthcare worker protection.
2. Reports, guidelines, and journal articles related to healthcare worker protection during the COVID-19 pandemic response were published/released between March 2020 (when COVID-19 was declared a global health threat and Kenya reported her first case) and March 2021.
3. Media articles were included in the review if they were published between March 2020 and March 2021.

Overall, 44 policy documents (legislation, public sector and health sector policy, COVID-19 related guidelines and reports) and 22 media articles were included in this review.

## RESULTS

### Governance of HCW Protections

#### Role of national and county governments in HCWs protections

HCW protection in Kenya is enshrined in the Constitution, specific legislation, regulations and guidance (Table 3).

**Table 3:** Provision for HCW protection drawn from legislation and policies.

| **Legislation** | **Provision for HCW protection** |
| --- | --- |
| The Constitution, 2010 | Article 41: ‘Every person has the right to fair labour practices. These include the right to; fair remuneration, reasonable working conditions, join and participating in trade union activities (except the Police and Defence Forces) which includes participating in a lawful strike (Article 41) (8). |
| Employment Act, 2007 | Requires employers (both public and private sectors), to provide medical treatment for employees including transport to and from the hospital at the employer’s cost unless provided free by the Government (Section 46((1);<br>Protects employees against dismissal for participating in trade union activities including strikes (46) (d)(i) (9). |
| Work Injury Benefits Act, 2007 | Requires public and private sector employers to obtain and maintain an insurance policy, to cover any liability that the employer may incur under this Act to any of his employees; (Section 38 (1)<br>Confers an employee with entitlements to compensation if the employee contracts a disease that arose out of and during the employee’s employment (Section 38(b) |
| Occupational Safety and Health Act, 2007 | Requires:<br>-Occupiers to ensure the safety, health, and welfare at work of all persons working in their workplace. (Section 6)<br>-Employer to maintain a register of all occupational diseases/injuries (Section 21 (2)<br>-Employer to provide protective personal equipment at the workplace<br>-Medical surveillance which includes planned periodic examination of employees, through clinical examination or medical tests of employees at the start, during and at the exit of employment (Section 2). |
| County Public Service and Human Resource Manual, 2013. | -Identifies the county government as an employer with the responsibility to provide medical cover to an employee and his family (pg. 48)<br>-Establishes a consultative committee consisting of up to three members from the County Government and three members from employees’ unions. The main functions of the Committee are to provide a platform for negotiation and secure the greatest cooperation between employers and employees (pg. 60)<br>-Chief Officer has a responsibility to ensure a safe working environment, provision of PPE to those working in hazardous working environments, medical examination of those working in hazardous environments (pg. 63)<br>-Provision to make claims for workplace injuries and diseases acquired in the course of employment (pg. 64) |
| Public Service and Human Resources Manual, 2016. | -Has provision for medical benefits for government employees and their dependents including medical ex-gratia assistance, where an employee exceeds in-patient medical cover (pg. 58-59)<br>-Requires all state departments and their agencies to have a health and safety committee (pg. 96) |
|  | -Requires employees to be provided with personal protective equipment (pg. 98);<br>-Highlights compensation for occupational disease, injury or death arising out of the course of employment (pg. 98)<br>-Identifies the National Treasury as the source of funds for compensation for work injuries and occupational diseases. (pp 101) |
| Healthcare Waste Management Strategic Plan 2015-2020 | -Establishes technical teams to guide Healthcare waste management to national and county governments (pg. 34)<br>-Identifies facility level (Infection Prevention and Control (IPC) committees as the structures that guide Healthcare waste management (pg. 20) |

Document reviews suggested that both the Public Service and Human Resource Policies and Procedures Manual (10) and the County Public Service and Human Resource Manual (11) are in use for the management of public sector healthcare workers at the national and county level respectively (12, 13). However, the county-level guidance did not address key Occupational Safety and Health (OSH) structures and processes such as OSH Committees at workplaces; funding sources, or funds and processes for accessing employee medical benefits even though these are included in the national level guidance (11, 14).

In practice, governance arrangements for HCW protection are shared by the national and county governments (Figure 2). Our review also highlighted an important role for non-state actors including labour unions, civil society organizations, and the media.

**Figure 2:**
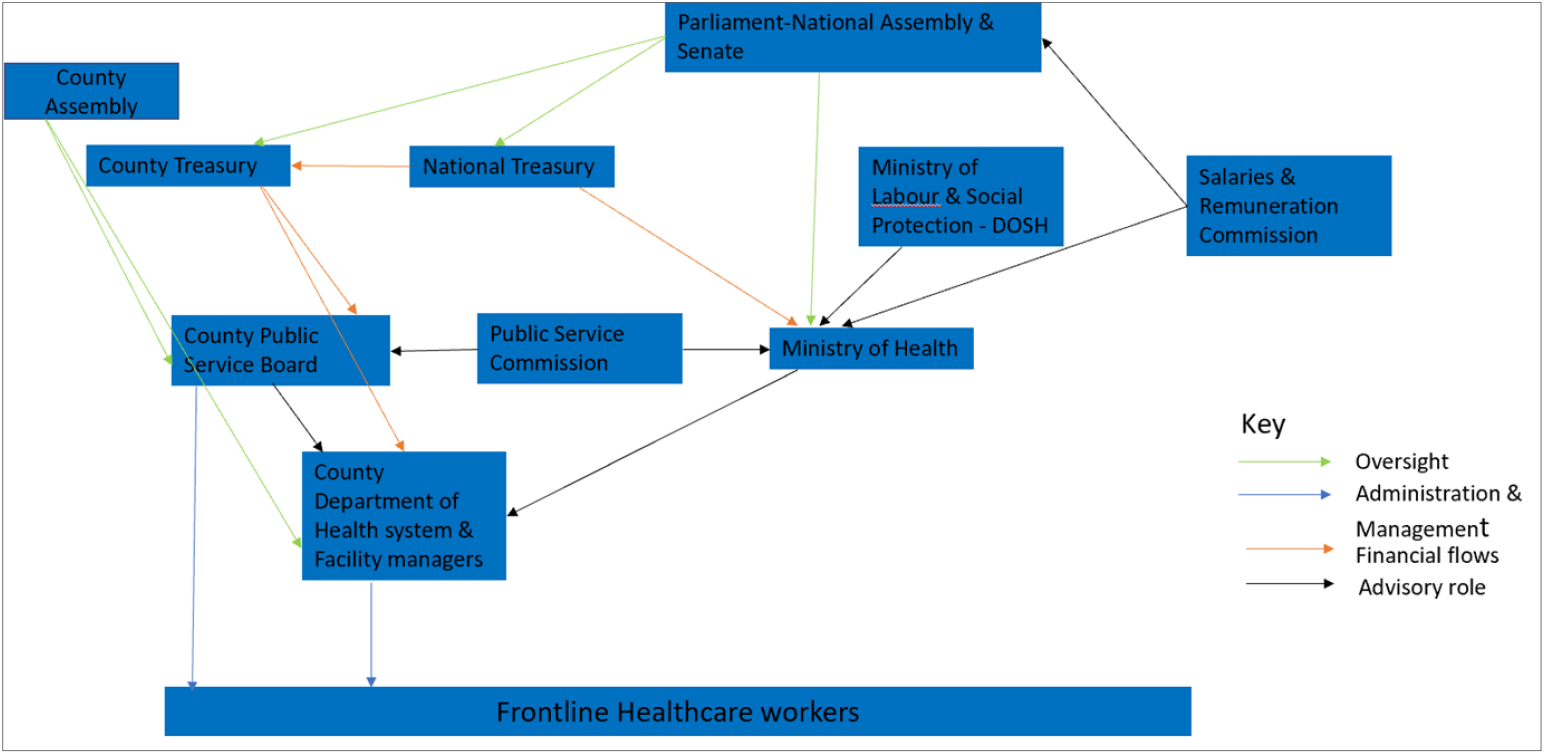
Governance architecture for HRH protection Source: World Bank

At the national level, the Public Service Commission (PSC) has delegated the role of Human Resources for Health (HRH) management to the MoH’s Directorate of Human Resources (15). The Directorate is responsible for the development of HRH policies and capacity building of counties for HRH management. MoH’s semi-autonomous agencies such as national referral hospitals have human resource management units within their organizational structures.

At the county level, HRH management is a joint responsibility of the County Public Services Board (CPSB) and the County Department of Health (CDoH). The CPSB has overall oversight and management of all the county public service employees (11). The county executive committee (CEC) member for health heads the CDOH and is responsible for the management and coordination of the county administration and its departments including the development and implementation of county policies. Every department including the County Department of Health (CDoH) has a Chief Officer who reports upwards to the CEC and Governor and is responsible for efficient management, administration and utilization of resources and public funds. The Chief Officer works closely with the County Director of Health and the County Health Management Team (CHMT) which comprises technical managers who provide support supervision and management for the sub-county levels and the Sub-County Health Management Team (SCHMT). Some counties have an HRH unit in keeping with MoH recommendations (16) whose function is to provide input on salary and benefits administration, be responsible for county-level HRH record management, disseminate HR policies and guidelines for the health sector, manage disputes between healthcare workers and county governments and ensure that healthcare workers have a safe, clean, healthy working environment (17). The unit is headed by an HRH manager who reports to the Chief Officer of Health.

Private sector HCW protections are provided through national legislation and regulation, and internal HR policies of the respective private sector entity. Additional oversight is through the Ministry of Labour and Social Protections’ (MoLSP) Directorate of Occupational Safety and Health Services (DOSHS). MoH guidance on staff qualifications and requirements for the performance of certain services or procedures also influence HWC protections in the private sector.

### Government measures enacted towards HCW protections

The government adopted several measures to ensure healthcare worker protection during the COVID-19 pandemic. We classified these using the lens of the TAPIC framework and identified their influence on HCW protection (Table 4).

**Table 4:**
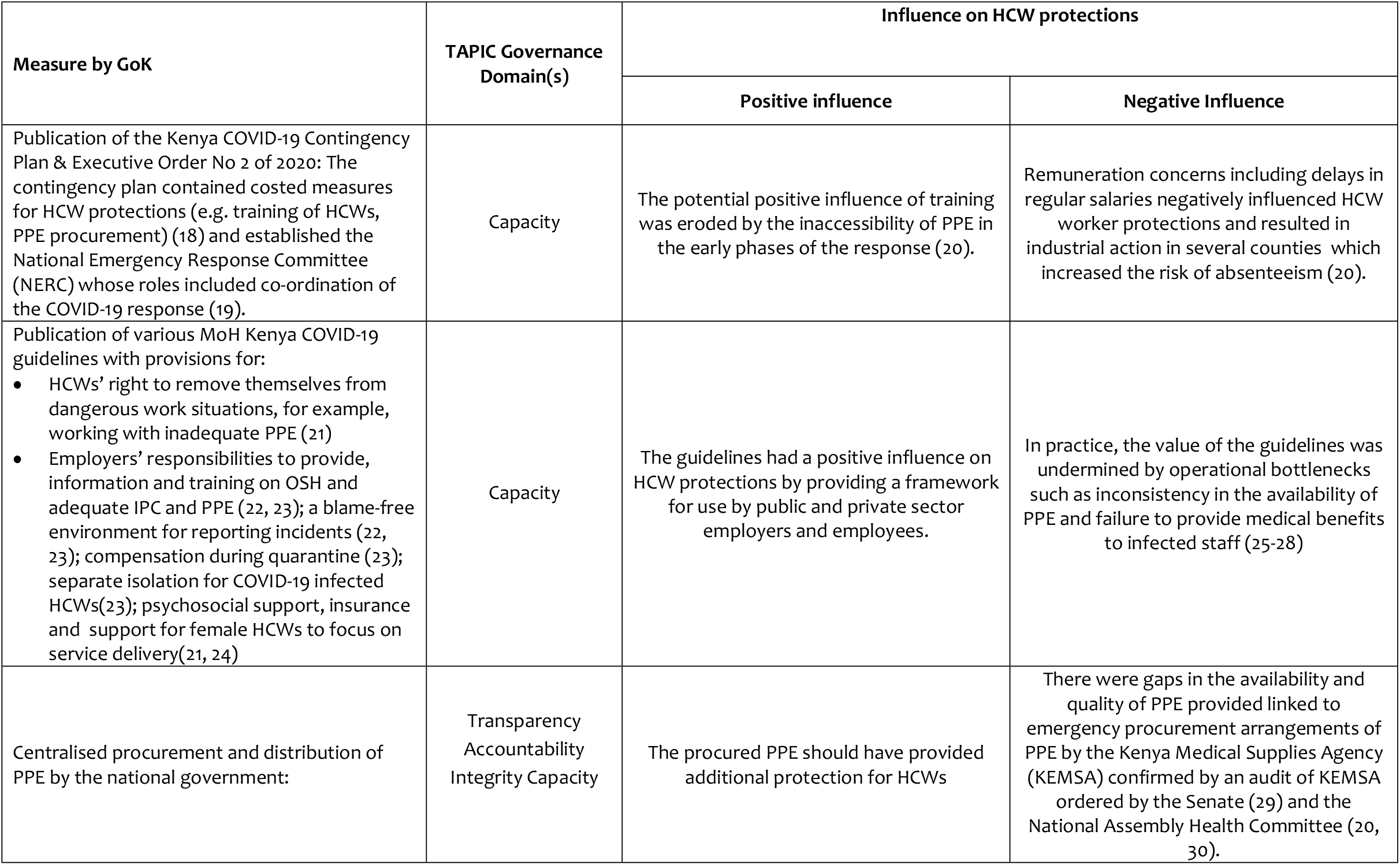
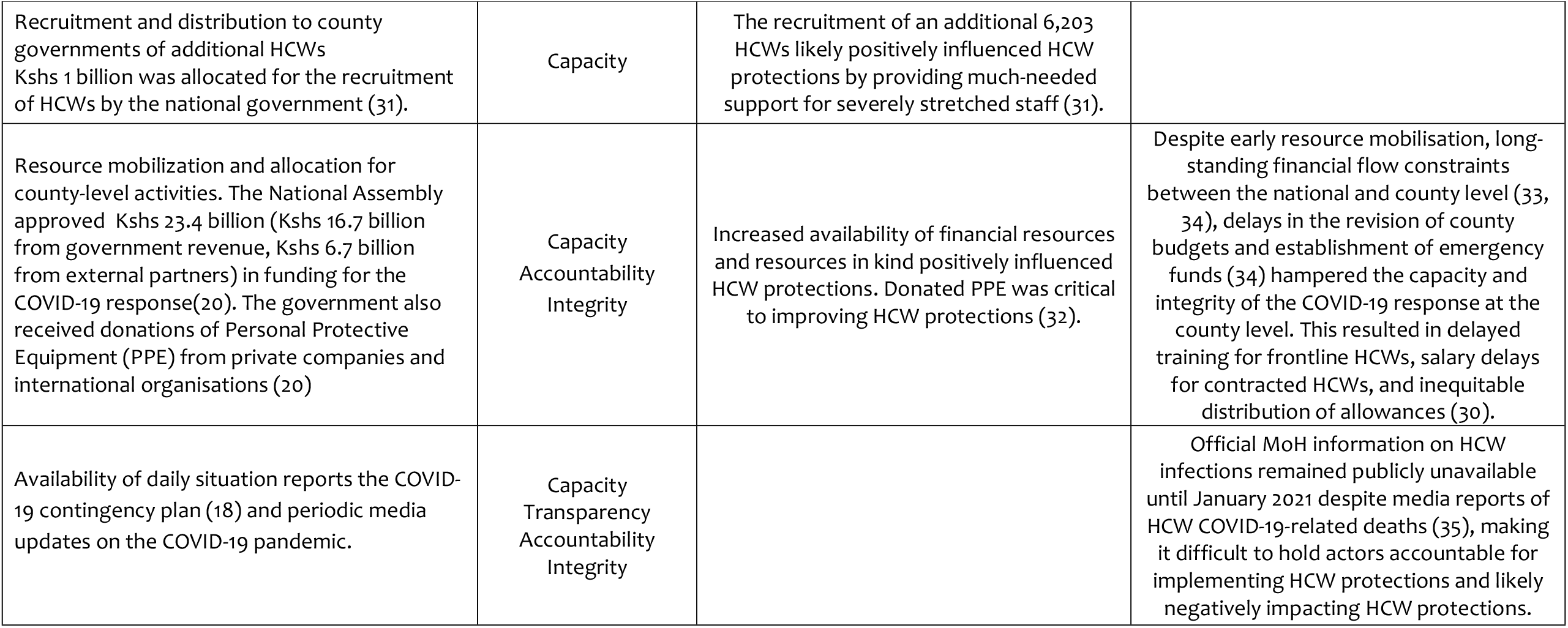
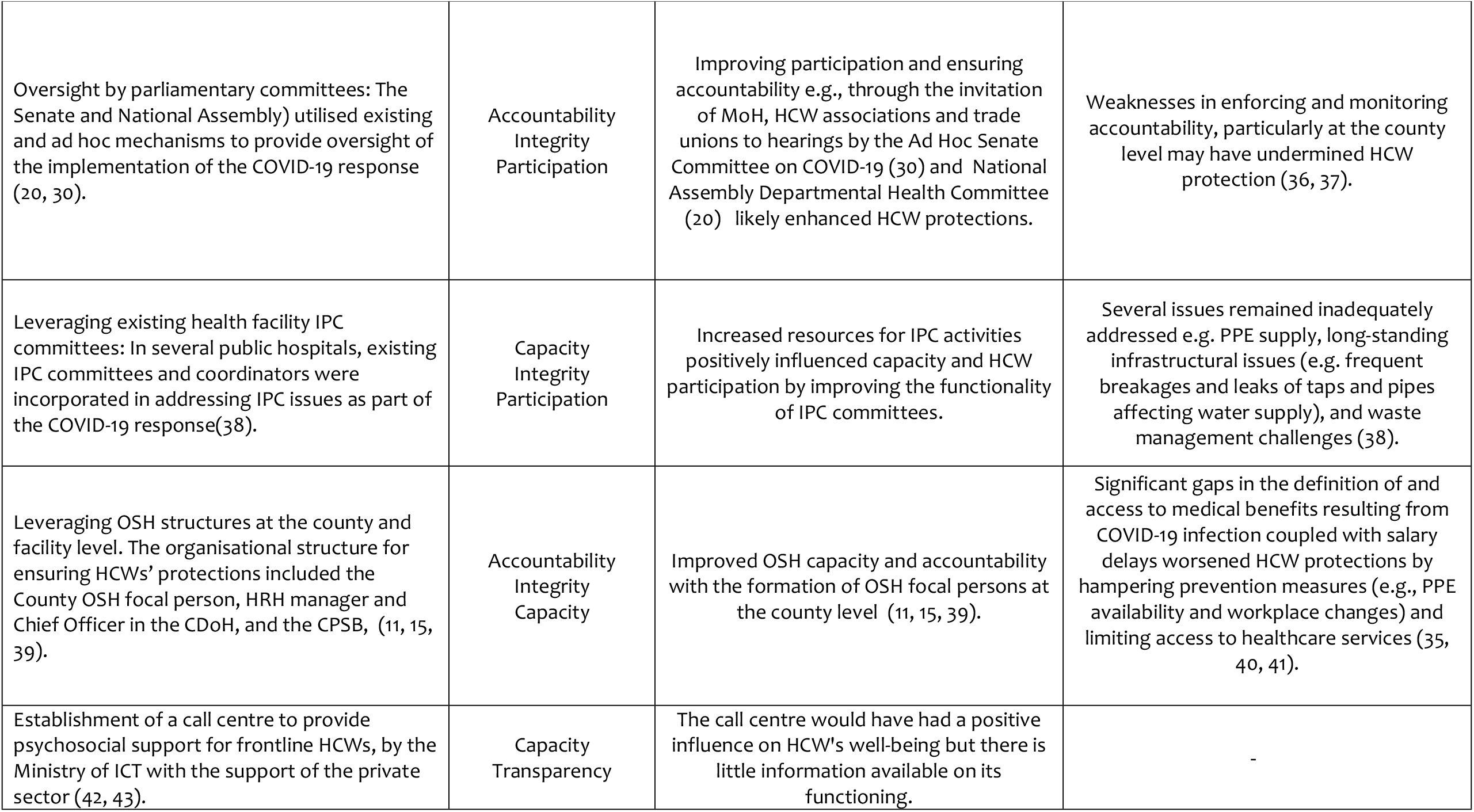
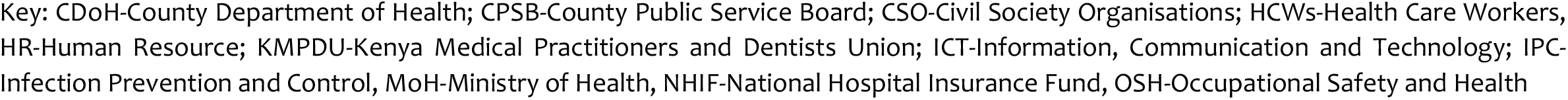
Summary of analysis of government interventions related to HCW protections

| Measure by GoK | TAPIC Governance Domain(s) | Influence on HCW protections |  |
| --- | --- | --- | --- |
|  |  | Positive influence | Negative Influence |
| Publication of the Kenya COVID-19 Contingency Plan & Executive Order No 2 of 2020: The contingency plan contained costed measures for HCW protections (e.g. training of HCWs, PPE procurement) (18) and established the National Emergency Response Committee (NERC) whose roles included co-ordination of the COVID-19 response (19). | Capacity | The potential positive influence of training was eroded by the inaccessibility of PPE in the early phases of the response (20). | Remuneration concerns including delays in regular salaries negatively influenced HCW worker protections and resulted in industrial action in several counties which increased the risk of absenteeism (20). |
| Publication of various MoH Kenya COVID-19 guidelines with provisions for:<br>• HCWs' right to remove themselves from dangerous work situations, for example, working with inadequate PPE (21)<br>• Employers' responsibilities to provide, information and training on OSH and adequate IPC and PPE (22, 23); a blame-free environment for reporting incidents (22, 23); compensation during quarantine (23); separate isolation for COVID-19 infected HCWs(23); psychosocial support, insurance and support for female HCWs to focus on service delivery(21, 24) | Capacity | The guidelines had a positive influence on HCW protections by providing a framework for use by public and private sector employers and employees. | In practice, the value of the guidelines was undermined by operational bottlenecks such as inconsistency in the availability of PPE and failure to provide medical benefits to infected staff (25-28) |
| Centralised procurement and distribution of PPE by the national government: | Transparency<br>Accountability<br>Integrity Capacity | The procured PPE should have provided additional protection for HCWs | There were gaps in the availability and quality of PPE provided linked to emergency procurement arrangements of PPE by the Kenya Medical Supplies Agency (KEMSA) confirmed by an audit of KEMSA ordered by the Senate (29) and the National Assembly Health Committee (20, 30). |
|  |  | Positive influence | Negative Influence |
| Recruitment and distribution to county governments of additional HCWs<br>Kshs 1 billion was allocated for the recruitment of HCWs by the national government (31). | Capacity | The recruitment of an additional 6,203 HCWs likely positively influenced HCW protections by providing much-needed support for severely stretched staff (31). |  |
| Resource mobilization and allocation for county-level activities. The National Assembly approved Kshs 23.4 billion (Kshs 16.7 billion from government revenue, Kshs 6.7 billion from external partners) in funding for the COVID-19 response(20). The government also received donations of Personal Protective Equipment (PPE) from private companies and international organisations (20) | Capacity<br>Accountability<br>Integrity | Increased availability of financial resources and resources in kind positively influenced HCW protections. Donated PPE was critical to improving HCW protections (32). | Despite early resource mobilisation, long-standing financial flow constraints between the national and county level (33, 34), delays in the revision of county budgets and establishment of emergency funds (34) hampered the capacity and integrity of the COVID-19 response at the county level. This resulted in delayed training for frontline HCWs, salary delays for contracted HCWs, and inequitable distribution of allowances (30). |
| Availability of daily situation reports the COVID-19 contingency plan (18) and periodic media updates on the COVID-19 pandemic. | Capacity<br>Transparency<br>Accountability<br>Integrity |  | Official MoH information on HCW infections remained publicly unavailable until January 2021 despite media reports of HCW COVID-19-related deaths (35), making it difficult to hold actors accountable for implementing HCW protections and likely negatively impacting HCW protections. |
|  |  | Positive influence | Negative Influence |
| Oversight by parliamentary committees: The Senate and National Assembly) utilised existing and ad hoc mechanisms to provide oversight of the implementation of the COVID-19 response (20, 30). | Accountability<br>Integrity<br>Participation | Improving participation and ensuring accountability e.g., through the invitation of MoH, HCW associations and trade unions to hearings by the Ad Hoc Senate Committee on COVID-19 (30) and National Assembly Departmental Health Committee (20) likely enhanced HCW protections. | Weaknesses in enforcing and monitoring accountability, particularly at the county level may have undermined HCW protection (36, 37). |
| Leveraging existing health facility IPC committees: In several public hospitals, existing IPC committees and coordinators were incorporated in addressing IPC issues as part of the COVID-19 response(38). | Capacity<br>Integrity<br>Participation | Increased resources for IPC activities positively influenced capacity and HCW participation by improving the functionality of IPC committees. | Several issues remained inadequately addressed e.g. PPE supply, long-standing infrastructural issues (e.g. frequent breakages and leaks of taps and pipes affecting water supply), and waste management challenges (38). |
| Leveraging OSH structures at the county and facility level. The organisational structure for ensuring HCWs' protections included the County OSH focal person, HRH manager and Chief Officer in the CDoH, and the CPSB, (11, 15, 39). | Accountability<br>Integrity<br>Capacity | Improved OSH capacity and accountability with the formation of OSH focal persons at the county level (11, 15, 39). | Significant gaps in the definition of and access to medical benefits resulting from COVID-19 infection coupled with salary delays worsened HCW protections by hampering prevention measures (e.g., PPE availability and workplace changes) and limiting access to healthcare services (35, 40, 41). |
| Establishment of a call centre to provide psychosocial support for frontline HCWs, by the Ministry of ICT with the support of the private sector (42, 43). | Capacity<br>Transparency | The call centre would have had a positive influence on HCW's well-being but there is little information available on its functioning. | - |

Key: CDoH-County Department of Health; CPSB-County Public Service Board; CSO-Civil Society Organisations; HCWs-Health Care Workers, HR-Human Resource; KMPDU-Kenya Medical Practitioners and Dentists Union; ICT-Information, Communication and Technology; IPC- Infection Prevention and Control, MoH-Ministry of Health, NHIF-National Hospital Insurance Fund, OSH-Occupational Safety and Health

### Experiences of HCWs in the private sector

An analysis of the reviewed documents provided little information on the experiences of HCWs in the private sector. Few private facilities, mainly large private hospitals within urban areas, provided COVID-19 care and treatment at the time of analysis (44). The limited engagement of private facilities in managing COVID-19 patients was linked to inadequate PPE and supplies, and an expectation that all suspected COVID-19 would be referred to public health facilities (44). Despite the lack of attention to the experiences of HCWs in the private sector, data from the KELIN studies suggest that in the early days of the COVID-19 response, there was low preparedness and responsiveness to HCW infections across both public and private sectors (26-28). These findings are however limited by the fact only 16% of 601 clinical officers, 17% of 155 nurses and 6% of 85 doctors (medical officers, pharmacists and dentists) surveyed worked in the private sector.

### Role of non-state actors

As early as April 2020, two healthcare worker unions, Kenya National Union of Nurses (KNUN) and Kenya Medical Practitioners, Pharmacists and Dentists Union (KMPDU) highlighted the inadequacy of PPE and lack of isolation centres for HCWs as key gaps in HCW protections (25). Professional associations such as the Kenya Medical Association (KMA), Kenya Dentists Association (KDA) and National Nurses Association of Kenya (NNAK) also issued recommendations for employers to ensure PPE provision, withdraw vulnerable HCWs from frontline roles, and advised HCWs of their right to decline to attend to suspected or confirmed COVID-19 cases if PPE was inadequate (45-47).

Civil society groups also played a prominent role in supporting the effective governance of HCW protections. Studies conducted in April 2020 by KELIN working with NNAK, KUCO and KMPDU demonstrated low levels of preparedness and response to HCW protections across various domains in both public and private sectors (26-28). The studies found that between 81-93% of HCWs lacked work injuries benefit cover; 72-82% had re-used PPE and 82-95% wanted psychosocial support.

The media also raised transparency and accountability questions through reports on the disappearance of donated PPE and other commodities (32), and poor reporting and under-utilisation of COVID-19 funds at the county level (48, 49). Increasing HCW agitation triggered by increasing COVID-19 infections and related deaths and perceived unresponsiveness of public employers escalated to county-level HCW strikes (50) and later to nationwide HCW strikes in December 2020 (35, 40, 41, 51, 52).

## DISCUSSION

This paper presents evidence of the governance arrangements for HCW protection in Kenya covering the first three waves of the COVID-19 pandemic in Kenya. Our review of governance capacities using the TAPIC framework found gaps in the design and implementation of measures to ensure HCW protection during the COVID-19 pandemic. These gaps likely contributed to the reported COVID-19 infection rates among HCWs that could have been avoidable.

Our analysis of the governance arrangements of HCW protections highlighted significant design gaps. Legislation concerned with employee protections did not align with decentralization arrangements, the evolution of workplaces away from industries, or the reality of emerging infectious diseases and public health emergencies (53-55). More recent policy documents, such as the MOH guidelines for OSH and National OSH policy have tried to catch up with decentralisation by including county governments as complementary actors in the delivery of HCW protection (39, 56). However, the translation of this policy to practice is constrained by insufficient monitoring and few active county OSH officers (57). Further, exposure to COVID-19 was identified as an occupational hazard for HCWs (58), a positive step in enhancing transparency and accountability during the COVID-19 pandemic, and in OSH practice for HCWs more broadly.

We also identified challenges in the implementation of HCW protections. We identified (i) fragmentation of responsibilities for OSH across multiple departments and levels of government (59, 60); (ii) inadequate operationalization of key provisions such as a medical scheme for employees; and (iii) insufficient surveillance and monitoring of OSH hazards and injuries among HCWs. Challenges with reporting of HCW OSH have been reported in other settings and there has been a call to countries to not only track HCW infections, deaths and other related occupational hazards, but also to make such information publicly accessible (61, 62).

The broader challenge of HCW information management has implications for preparedness and response. Poor information management hampered response in Kenya and other settings with challenges in confirming access to PPE (63). Better management of information would support the real-time implementation of response measures such as increasing access to PPE, supporting effective resource reallocation, and supporting the rapid investigation of measures such as vaccination as demonstrated in the current pandemic (64).

Several pre-pandemic contextual factors may also have contributed to the observed shortcomings in HCW protections. Kenya scored poorly on the coordination of public health emergency preparedness and response in the Joint Inspection Report on International Health Regulations Conducted in 2015 (65). Coordination challenges between national and county-level health system actors (66, 67) may also have exacerbated the usual difficulties of top-down implementation approaches such as poor information flow and incentive misalignment (68, 69). Chronic underinvestment (70) and inefficiencies (71) in the health sector also meant that the system was less well-prepared to respond to increased demands for HCW protection. This is despite recent assessments showing the value of an investment in HCW protection through measures such as PPE (63). The establishment of the National Public Health Institute to coordinate public health emergency preparedness and response should go some way in bridging this gap (63).

### Study limitations and Strengths

Our findings should be interpreted in light of the following limitations. First, the scoping review was limited by a scarcity of documents that adopted a broad definition of HCWs. The majority of the reviewed study documents focused on three cadres (nurses, clinical officers, and doctors) with little attention to other cadres and non-clinical staff. Second, few documents reported on HCW experiences in the private sector even though the private sector accounts for approximately 50% of health service provision in Kenya (72) (and therefore employs a significant proportion of HCWs). Third, interviews with key informants could have complemented the findings from the study. Despite these limitations, the article has several strengths. First, it explored legislation and policy guidelines and considered proposed policy with findings from government reports and media accounts. This broad-based review enriches the body of knowledge on HCW protection status and needs. Second, the application of a TAPIC framework allowed for an objective discussion of policy and practice and has enabled reflection on the extent to which HCW protections proposed and implemented during the early days of the COVID-19 pandemic met HCW’s needs.

## CONCLUSIONS

The study revealed how implementation factors such as capacity, transparency, and accountability gaps interacted with policy design gaps to undermine HCW protection. Existing laws and policies promoting HCW protections are a strength that Kenya can build on for greater responsiveness to HCWs’ protections even beyond the COVID-19 pandemic. Consequently, it is paramount to (i) update relevant labour relations and OSH legislation and policies to reflect devolved governance and identify infectious diseases as workplace hazards for HCWs; (ii) enhance the functionality of operational structures for HCW occupational safety and health during emergencies specifically through legal and administrative provisions for national-level coordination and implementation; (iii) strengthen information management around HCW safety and health including a broader definition of HCWs and a broader view of health including mental health; (iv) develop a plan to institutionalize financial and health system responsiveness during public health emergencies e.g., to account for fast-tracked procurement procedures or funds flows; and (v) develop a costed plan for the preparedness and response measures for HCW protection during public health emergencies e.g., to account for access to PPE and emerging measures such as COVID-19 vaccination.

## Data Availability

This study involved a scooping review rather than primary data collection. All reviewed documents have been referenced in the manuscript.

## Ethics

Not applicable. The study utilised secondary sources of data.

